# Effectiveness of a nation-wide COVID-19 vaccination program in Mexico

**DOI:** 10.1101/2022.04.04.22273330

**Authors:** Omar Yaxmehen Bello-Chavolla, Neftali Eduardo Antonio-Villa, Sergio Iván Valdés-Ferrer, Carlos A. Fermín-Martínez, Luisa Fernández-Chirino, Daniel Ramírez-García, Javier Mancilla-Galindo, Ashuin Kammar-García, José Alberto Ávila-Funes, Clemente Humberto Zúñiga-Gil, Miguel García-Grimshaw, Santa Elizabeth Ceballos-Liceaga, Guillermo Carbajal-Sandoval, José Antonio Montes-González, Christian Arturo Zaragoza-Jiménez, Gabriel García-Rodríguez, Ricardo Cortés-Alcalá, Gustavo Reyes-Terán, Hugo López-Gatell, Luis Miguel Gutiérrez-Robledo

## Abstract

**BACKGROUND:** Vaccination has been effective in ameliorating the impact of COVID-19. However, estimation of vaccine effectiveness (VE) is still unavailable for some widely used vaccines and underrepresented groups. Here, we report on the effectiveness of a nation-wide COVID-19 vaccination program in Mexico.

**METHODS:** We used a test-negative design within a national COVID-19 surveillance system to assess VE of the BNT162b2, mRNA-12732, Gam-COVID-Vac, Ad5-nCoV, Ad26.COV2.S, ChAdOx1 and CoronaVac vaccines, against SARS-CoV-2 infection, COVID-19 related hospitalization and death for adults ≥18 years in Mexico. VE was estimated using Cox proportional hazard models considering time-varying vaccination status in partial and fully vaccinated individuals compared to unvaccinated adults, adjusted by age, sex, comorbidities and municipality. We also estimated VE for adults ≥60 years, for cases with diabetes and comparing periods with predominance of variants B.1.1.519 and B.1.617.2.

**RESULTS:** We assessed 793,487 vaccinated compared to 4,792,338 unvaccinated adults between December 24^th^, 2020, and September 27^th^, 2021. VE against SARS-CoV-2 infection was highest for fully vaccinated individuals with mRNA-12732 (91.5%, 95%CI 90.3-92.4) and Ad26.COV2.S (82.2%, 95%CI 81.4-82.9), whereas for COVID-19 related hospitalization were BNT162b2 (84.3%, 95%CI 83.6-84.9) and Gam-COVID-Vac (81.4% 95%CI 79.5-83.1) and for mortality BNT162b2 (89.8%, 95%CI 89.2-90.2) and mRNA-12732 (93.5%, 95%CI 86.0-97.0). VE for all evaluated vaccines was reduced for adults ≥60 years, people with diabetes, and in periods of Delta variant predominance.

**CONCLUSIONS:** All evaluated vaccines were effective against SARS-CoV-2 infection and COVID-19 related hospitalization and death. Mass vaccination campaigns with multiple vaccine products are feasible and effective to maximize vaccination coverage.

## INTRODUCTION

Coronavirus Disease (COVID-19) has imposed a significant public health challenge worldwide. In Mexico, mitigation of COVID-19 has imposed a significant challenge due to the large impact of cardio-metabolic diseases and long-standing socio-demographic and healthcare inequalities in the region^1–3^. The most significant contributing factor to decreasing the impact of COVID-19 has been the availability of vaccines to prevent symptomatic and severe forms of severe acute respiratory syndrome coronavirus 2 (SARS-CoV-2) infections^4,5^. Available COVID-19 vaccines are effective in preventing symptomatic SARS-CoV-2 infection and COVID-19 related hospitalization and death^6–12^. Despite the availability or randomized clinical trials to inform vaccine efficacy, population-based studies to estimate vaccine effectiveness are required to evaluate and inform future vaccine rollouts, assess and prioritize eligibility for vaccine boosters and reduce vaccine hesitancy^13^.

The COVID-19 vaccination program in Mexico sequentially implemented seven nationally available vaccines: BNT162b2, mRNA-12732, Gam-COVID-Vac, Ad5-nCoV, Ad26.COV2.S, ChAdOx1, and CoronaVac. To date, population-based studies have reported population-wide effectiveness of the BT162b2 vaccine^14–16^, ChAdOx1^17–19^, mRNA-12732^20,21^, Ad26.COV2.S^22,23^, and the rAd26 component of the Gam-COVID-Vac vaccine^24^. Nevertheless, studies in Latin America have been scarce, with only two reports on the efficacy of the CoronaVac^25,26^ vaccine. The Mexican COVID-19 vaccination program offers a unique opportunity to evaluate the simultaneous effectiveness and impact of a diverse set of available COVID-19 vaccines against SARS-CoV-2 infection and COVID-19 related hospitalization and death, and report estimates for effectiveness of the Gam-COVID-Vac and Ad5-nCoV vaccines, which have had limited population-based evidence beyond clinical trials^7,12^. Furthermore, underrepresentation of older adults and patients with conditions which may lead to impaired immune response to the vaccine and who may benefit from prioritization for booster allocation, are often underrepresented in clinical trials; evaluating effectiveness for COVID-19 vaccines in these groups remains a public health priority^27^. In this study, we sought to provide the first nation-wide evaluation for the effectiveness of nationally available COVID-19 vaccines for SARS-CoV-2 infections and COVID-19 hospitalization and death, and the impact of circulating variants, older age and comorbidities in modifying effectiveness for COVID-19 vaccines implemented within the Mexican vaccination program during 2020-2021.

## METHODS

### Study population and design

We conducted a retrospective national cohort, using a test-negative design to evaluate vaccine effectiveness for nationally available COVID-19 vaccines in Mexico. We included adults aged 18 years or older, evaluated within the Sistema de Vigilancia Epidemiológica de Enfermedades Respiratorias (SISVER) database of the General Directorate of Epidemiology of the Mexican Ministry of Health. SISVER is a nation-wide sentinel surveillance system which collects daily updated clinical and epidemiological information of suspected COVID-19 cases in Mexico^1,2^. Vaccination status, date and specific vaccine product was collected from evaluated persons as part of epidemiological follow-up of suspected COVID-19 cases. For this analysis, we included cases evaluated in SISVER from December 24^th^, 2020, until September 27^th^, 2021. We compared cases with at least one dose of any available vaccine in Mexico with unvaccinated cases during the same period. As per WHO recommendations and given previous evidence on immune response to COVID-19 vaccination, we only included cases occurring ≥14 days after receiving at least one vaccine dose^26,27^.

### COVID-19 vaccination in Mexico

Mexico approached its vaccination strategy by incorporating multiple vaccine products to maximize national vaccination coverage. SARS-CoV-2 vaccines which were applied in Mexico during this period included BNT162b2, mRNA-12732, Gam-COVID-Vac, Ad5-nCoV, Ad26.COV2.S, ChAdOx1 and CoronaVac^28^. Vaccination protocols for most vaccines included a 2-dose regime as recommended by manufactures, with the exemption of 1-dose vaccines for Ad5-nCoV and Ad26.COV2.S. Fully vaccinated individuals were considered if they completed the vaccination protocol for two or one dose vaccines and partially vaccinated individuals were considered if they only completed one out of a two-dose vaccine protocol. All vaccines received emergency authorization for its use during the COVID-19 pandemic in Mexico by the Federal Commission for the Protection against Sanitary Risks (COFEPRIS), the Mexican health regulatory agency, after demonstrating efficacy in Phase III studies^29^. Vaccination rollout for population ≥18 years started for essential healthcare workers on December 24^th^, 2020 and continued with adults over 60 years and the rest of the population starting on in February 2021, with an estimated vaccine rollout of 44.63 million adults with a full vaccination protocol and 18.85 million with a partial vaccination protocol successfully completed as of September 27^th^, 2021, accounting for a coverage of 49% of the population with at least one dose^30^. Additional details on nationally available vaccine products in Mexico are reported in **Supplementary Material**.

### Outcomes

We calculated vaccine effectiveness (VE) based on three primary outcomes: 1) SARS-CoV-2 infections including both symptomatic and asymptomatic individuals, which considered laboratory confirmed SARS-CoV-2 infections with either rapid antigen test or RT-PCR. All diagnostic tests used to identify SARS-CoV-2 infections were certified by COFEPRIS and identified as per WHO recommendations^31^. 2) COVID-19 related hospitalization and 3) COVID-19 related death. We also further stratified VE based the following criteria:

1. *Predominant SARS-CoV-2 variant* – Relevant variants of interest in Mexico were screened by the Mexican Genomic Surveillance Consortium (COVIDGen-Mex)^32^. During the evaluated period, COVIDGen-Mex reported predominance of the B.1.1.519 variant prior to July 1^st^, 2021, and the B.1.617.2 (Delta) variant after this period. To evaluate changes in VE based on predominant variant circulation, we stratified the analyses based on date of symptom onset prior to July 1^st^, 2021, as B.1.1.519 predominance and after that period Delta predominance.
2. *Age strata* – Given underrepresentation of older adults in vaccine clinical trials and observed inequities on the impact of COVID-19 on this population in Mexico^2^, we evaluated VE using 60 years as the cut-off for age.
3. *Comorbidities* – Prevalence of diabetes and obesity in Mexican population are high and the impact of their impact on the course of the COVID-19 pandemic in Mexico have been widely reported^1,33,34^. To this end, we also evaluated vaccine effectiveness for individuals with diabetes and, secondarily, obesity.

### Statistical analyses

VE was estimated using a Cox proportional hazard regression model, which accounts for the time-varying covariate of time from beginning of follow-up until vaccination, as previously described^26,35^. The model is presented as follows:

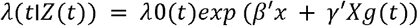

Where *λ*(*t*|*Z*(*t*)) is the baseline risk function for individuals at the beginning of the study without assumption of an underlying distribution, *β*′ represents a vector of fixed covariates and *γ*′*Xg*(*t*) represents the time-varying vaccination status for all evaluated persons. Exposure time was calculated from December 24^th^, 2020, until the onset of each evaluated outcome or until last follow-up, whichever occurred first. Vaccination status will be evaluated as a time-dependent covariate for each evaluated vaccine, calculated as the time from study entry to application of the first dose of any received vaccine. VE will be estimated for each evaluated vaccine product, where effectiveness is defined as 1 minus the hazard ratio of the resulting Cox model. For each comparison, each individual vaccine product was compared to unvaccinated population; this was also conducted for subgroup analyses for age, predominant circulating variant and comorbidities. We fitted all models for each vaccine product adjusted for age, sex, comorbidities and stratified per municipality of origin to account for region-level variation in pandemic dynamics, including all stratified analyses. All statistical analyses were conducted using R version 4.1.2.

## RESULTS

### Study population

We evaluated 793,487 vaccinated persons aged 18 or older and with ≥14 days from vaccination who were evaluated for suspected SARS-CoV-2 infection from December 24^th^, 2020, until September 27^th^, 2021. Overall, 437,968 cases were fully vaccinated with either one or two-dose regimes and 355,519 were partially vaccinated. The most frequent vaccines with at least one dose were ChAdOx1 (n=292,800), followed by BNT162b2 (n= 250,806), Ad5-nCoV (n= 87,585), Gam-COVID-Vac (n= 79,551), CoronaVac (n=62,474), Ad26.COV2.S (n=14,870), and mRNA-12732 (n=5,401). During the study period 4,792,338 unvaccinated individuals ≥18 years were assessed and included as controls in the study. Comparison of demographic characteristics and clinical outcomes disaggregated by vaccination status and type are presented in **Supplementary Materials**. Of interest, most vaccinated cases were in the 30-59 years age group. Regarding vaccine distribution the Ad26.COV2.S vaccine was used in a younger population, and the Gam-COVID-Vac and CoronaVac vaccines in a generally older population. Cases with diabetes more frequently reported vaccination with the BT162b2, Gam-COVID-Vac and the CoronaVac vaccines. A full flowchart of evaluated individuals and the distribution of vaccines within the SISVER registry and included in the study is provided in **Figure 1**.

**Figure 1.**
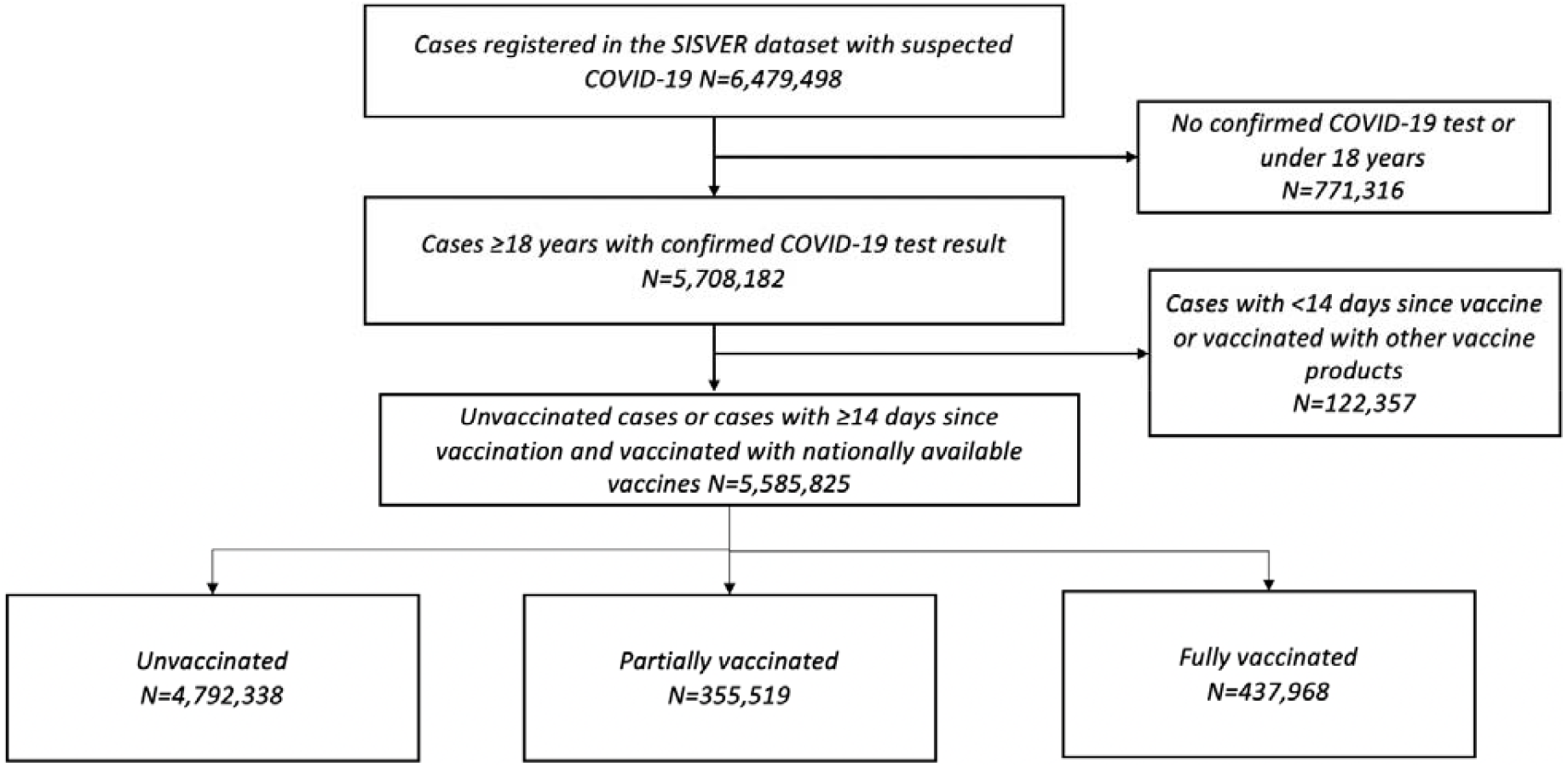
Flow diagram of individuals registered in the SISVER dataset from December 24^th^, 2020 until September 27^th^, 2021 after consideration of exclusion criteria for inclusion in our study.

### Incidence of COVID-19 and related outcomes during the study period

During the study period, a total of 1,700, 212 confirmed SARS-CoV-2 cases were reported. Overall, unvaccinated individuals accounted for 549,685,635 person-days of follow-up, with 80,621,913 person-days for partially vaccinated and 98,631,832 person-days for fully vaccinated individuals. Incidence rates for SARS-CoV-2 infection in these subgroups were 1.19 cases per 1,000 person-days for fully vaccinated individuals, followed by 1.41 cases per 1,000 person-days for partially vaccinated individuals and 2.67 cases per 1,000 person-days for unvaccinated individuals. We identified 230,780 COVID-19 related hospitalizations and 112,234 COVID-19 confirmed deaths. Cumulative incidence of SARS-CoV-2 infection and COVID-19 related hospitalizations and deaths were lower for vaccinated individuals for all evaluated vaccine products (**Figure 2**).

**Figure 2.**
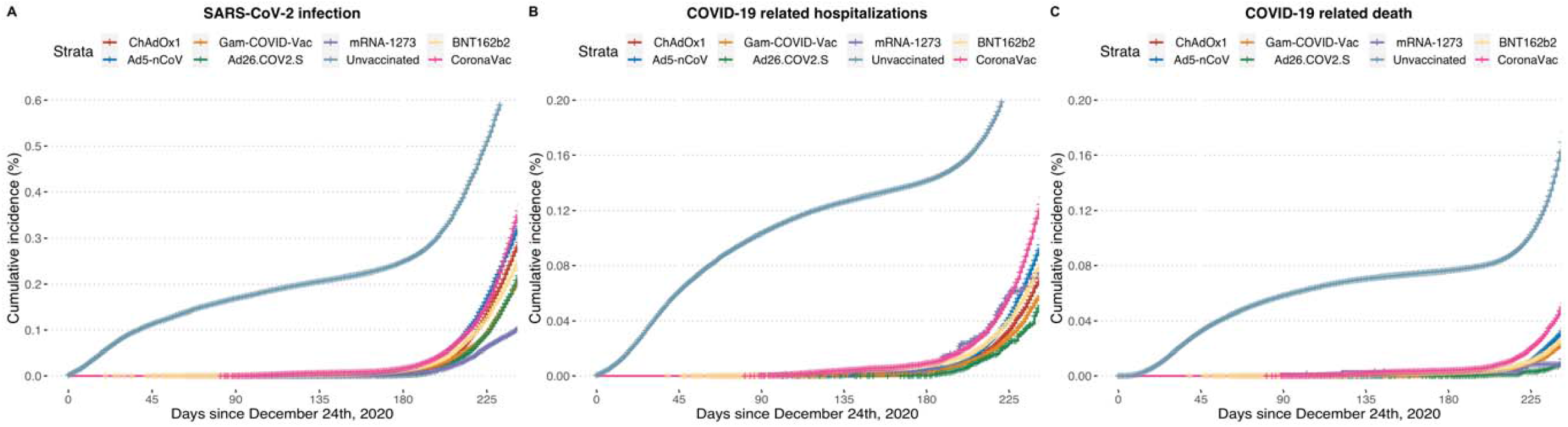
Cumulative incidence plots for laboratory confirmed SARS-CoV-2 infection, COVID-19 related hospitalization and death for unvaccinated individuals compared with individuals with at least one dose of all evaluated nationally available vaccines after the start of follow-up from December 24^th^, 2020.

### Vaccine effectiveness for nationally available 2-dose vaccines

Detailed effectiveness for SARS-CoV-2 infection and COVID-19 related hospitalization and mortality are available in **Table 1** and stratified by subgroups in **Figure 3**. For the BNT162b2 mRNA vaccine, effectiveness against SARS-CoV-2 infection in partially vaccinated individuals was 63.59% (95%CI 62.87-64.3) and increased with a 2-dose regime to 80.34% (95%CI 80.11-80.57). Protection for fully vaccinated individuals was 84.26% (95%CI 83.61-84.89) for hospitalization and 89.83% (95%CI 89.16-90.46) for mortality. In fully vaccinated individuals, decreased effectiveness for SARS-CoV-2 infection and COVID-19 related mortality was observed for individuals ≥60 years or those with diabetes. Protection against the predominant Delta variant was decreased for SARS-CoV-2 infection but slightly increased against COVID-19 related mortality compared to the period with predominance of the B.1.1.519 variant (**Supplementary Material**).

**Table 1.**
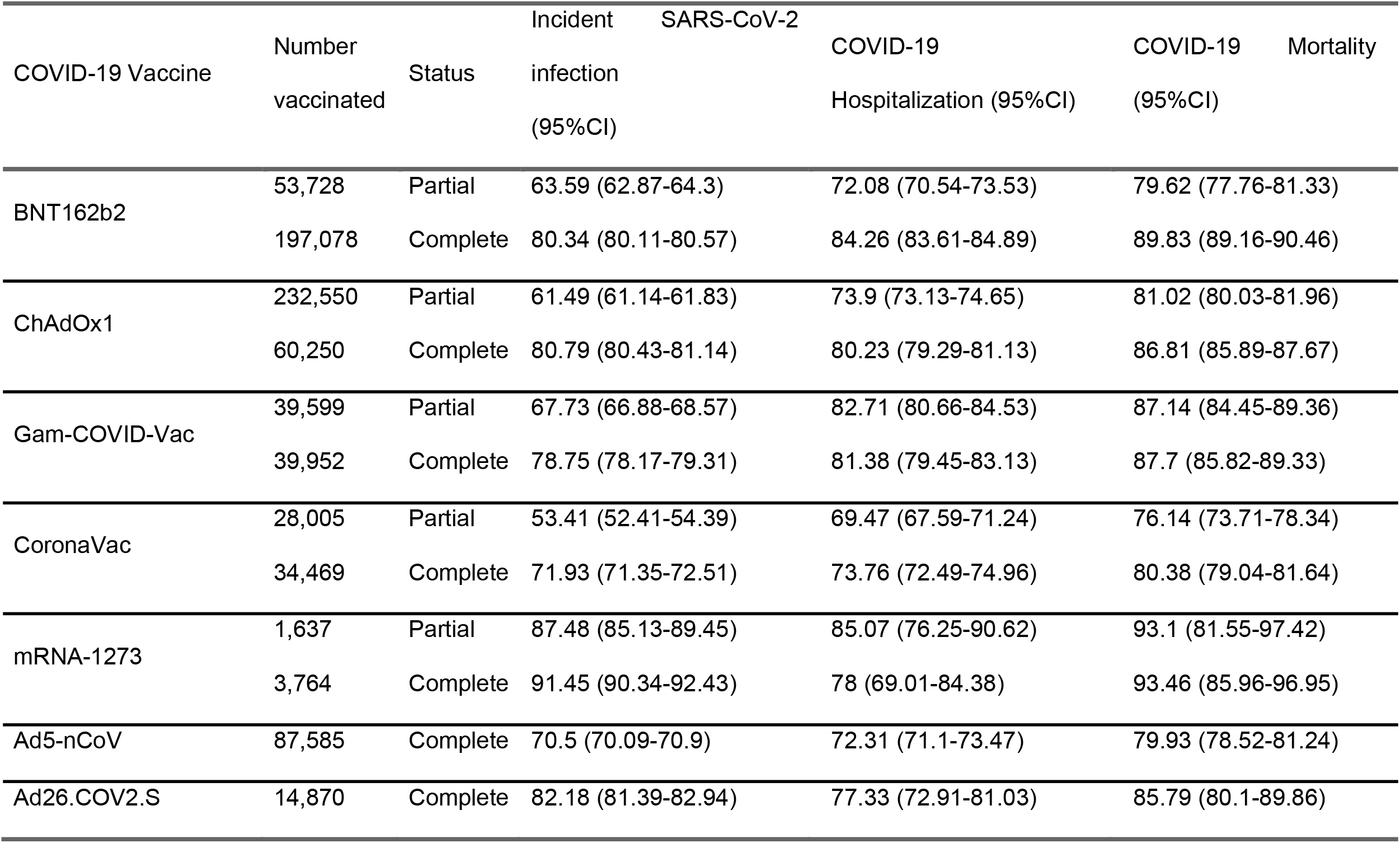
Estimated vaccine effectiveness for all nationally available COVID-19 vaccines in Mexico compared to unvaccinated adults (≥18 years) from December 24^th^, 2020, until September 27^th^, 2021. Effectiveness was estimated against laboratory-confirmed SARS-CoV-2 infection, COVID-19 related hospitalization and death. Partial and fully vaccinated status were considered for individuals with ≥14 days from vaccination until outcome.

**Figure 3.**
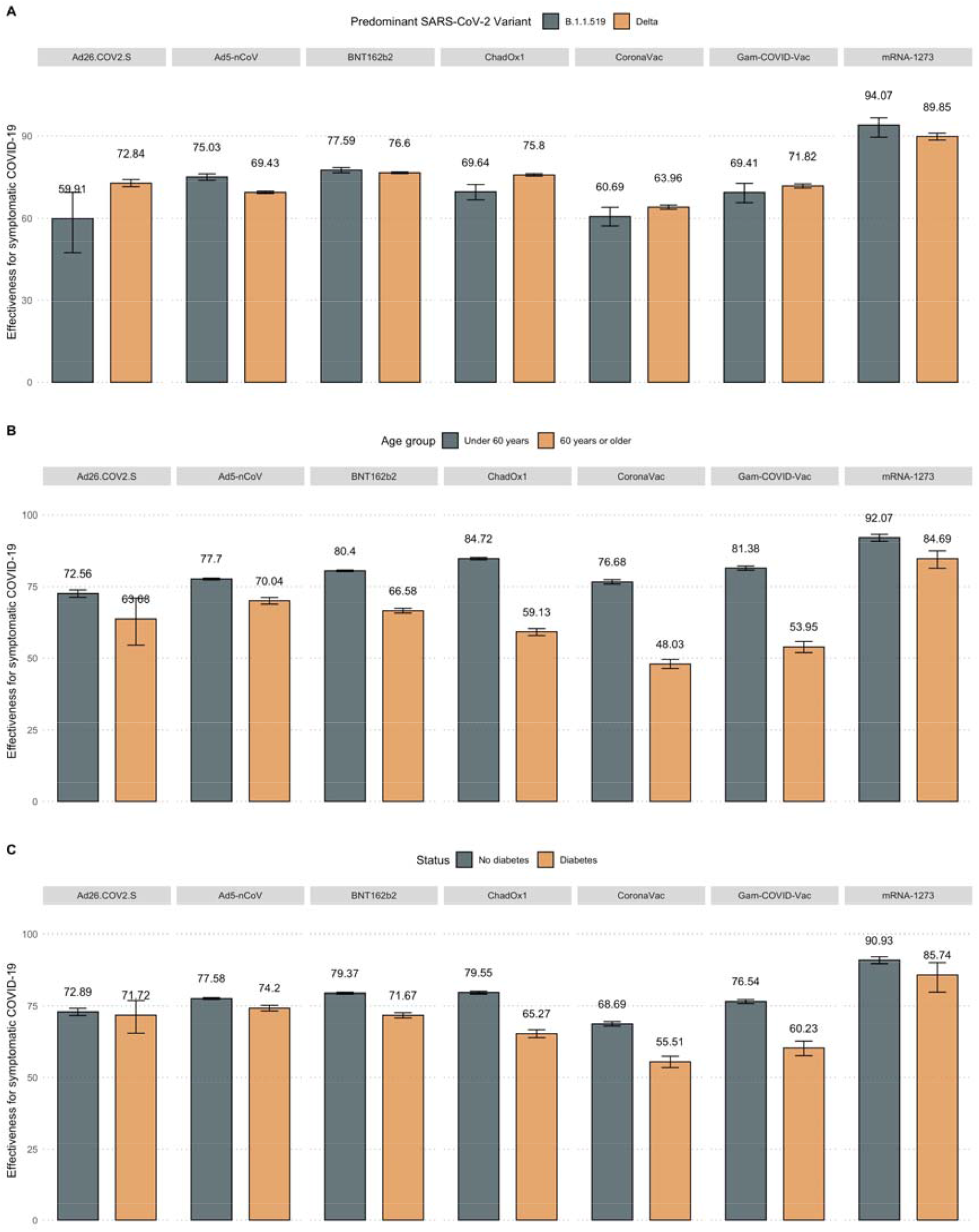
Estimated vaccine effectiveness against SARS-CoV-2 infection for nationally available COVID-19 vaccines stratified by predominant circulating SARS-CoV-2 variant (B.1.1.519 vs. B.1.617.2, A), age group (over vs. under 60 years, B) and diabetes status (C).

For the ChAdOx1 vaccine, effectiveness against SARS-CoV-2 infection in partially vaccinated individuals was 61.49% (95%CI 61.14-61.83), which increased in fully vaccinated individuals with a 2-dose regime to 80.79% (95%CI 80.43-81.14). Regarding hospitalization, the ChAdOx1 vaccine reached an effectiveness of 80.23% (95%CI 79.29-81.13) against COVID-19 related hospitalization and 86.81% (95%CI 85.89-87.67) for mortality. Lower effectiveness was observed for adults ≥60 years, and those with diabetes or obesity as well as during predominance of the Delta variant for both SARS-CoV-2 infection and COVID-19 related mortality (**Figure 3, Supplementary Material**).

The rAd26 first-dose component of the Gam-COVID-Vac vaccine reached an effectiveness of 67.73% (95%CI 66.88-68.57), against SARS-CoV-2 infection, and increased with the second rAd5 component to 78.75% (95%CI 78.17-79.31). Effectiveness for hospitalization in fully vaccinated individuals wash 81.38% (95%CI 79.45-83.13) and 87.7% (95%CI 85.82-89.33) for mortality. Effectiveness in fully vaccinated individuals with the Gam-COVID-Vac vaccine against SARS-CoV-2 infection markedly decreased for adults ≥60 years, and those with obesity or diabetes or during periods of Delta variant predominance (**Figure 3A**). Decreased effectiveness was also observed for COVID-19 mortality for these evaluated categories, except for periods of Delta predominance (**Supplementary Material**).

The inactivated SARS-CoV-2 CoronaVac vaccine reported an effectiveness of 53.41% (95%CI 52.41-54.39) for SARS-CoV-2 infection in partially vaccinated and 71.93% (95%CI 71.35-72.51) in fully vaccinated individuals. Protection against COVID-19 related hospitalization in fully vaccinated individuals was 73.76% (95%CI 72.49-74.96) and 80.38% (95%CI 79.04-81.64) for mortality. CoronaVac showed decreased effectiveness in adults >60 years, with obesity or those with diabetes for both SARS-CoV-2 infection and COVID-19 related death. Nevertheless, this vaccine did not display decreased effectiveness during periods of predominance of the Delta variant (**Figure 3A**).

Finally, for the mRNA-1273 vaccine, we observed the highest effectiveness against SARS-CoV-2 infection in partially vaccinated individuals with 87.48% (95%CI 85.13-89.45), which increased to 91.45% (95%CI 90.34-92.43) in fully vaccinated individuals. For protection against COVID-19 related hospitalization, the mRNA-1273 vaccine reached 78.0% (95%CI 69.01-84.38) effectiveness and 93.46% (95%CI 85.96-96.95) for mortality in fully vaccinated individuals. Decreased effectiveness was only observed for adults ≥60 years for SARS-CoV-2 infection, with stable effectiveness for all other evaluated categories (**Figure 3B, Supplementary Material)**.

### Vaccine effectiveness for nationally available 1-dose vaccines

Regarding the Ad5-nCoV vaccine, we observed an effectiveness for SARS-CoV-2 infection of 70.5% (95%CI 70.09-70.9), 72.31% (95%CI 71.1-73.47) for hospitalization and 79.93% (95%CI 78.52-81.24) for mortality (**Table 1**). Effectiveness against SARS-CoV-2 infection decreased for adults ≥60 years, with diabetes and predominance of the Delta variant (**Figure 3**); however, against COVID-19 related death this reduction was only observed for adults ≥60 years or cases with diabetes, with stable effectiveness during periods with predominance of the Delta compared with the B.1.1.519 variant (**Supplementary Material**).

Finally, the Ad26.COV2.S vaccine yielded the second highest effectiveness against SARS-CoV-2 infection with 82.18% (95%CI81.39-82.94), 77.33% (95%CI 72.91-81.03) for hospitalization and 85.79% (95%CI 80.1-89.86) for mortality. For SARS-CoV-2 infection this vaccine displayed stable effectiveness for all evaluated categories, with decreased effectiveness only for individuals with diabetes against COVID-19 related mortality.

## DISCUSSION

In this study of 793,487 vaccinated individuals ≥18 years compared with 4,792,338 unvaccinated controls, we provided estimates of the effectiveness of seven COVID-19 vaccines for the prevention of laboratory confirmed SARS-CoV-2 infection as well as COVID-19 related hospitalization and death, implemented within a national COVID-19 vaccination program in Mexico. Overall, all vaccines implemented in the Mexican National Vaccination Program proved to be effective in protecting against all evaluated outcomes in fully vaccinated adults; VE for SARS-CoV-2 infection was reduced in for adults ≥60 years, people living with diabetes and during periods with predominance of the Delta compared with the B.1.1.519 variants of SARS-CoV-2 in Mexico. Ours is the first study to additionally report estimated of effectiveness for the heterologous Gam-COVID-Vac and the Ad5-nCOV vaccines, which were widely used in Mexico to rapidly increase vaccine coverage, and which proved to be effective against all evaluated outcomes.

Vaccine effectiveness estimates in Mexico were similar to those reported in other countries and settings^22,24–26,35^, with similar reductions of effectiveness for periods of increased circulation of the Delta variant of SARS-CoV-2^36–38^. Notably, all estimates of vaccine effectiveness observed in our study coincide with the circulation of two main SARS-CoV-2 variants, the B.1.1.519 which was highly prevalent in Mexico from December 2020 until July 2021^39,40^ and the Delta variant (B.1.617.2)^41^. Estimates for VE for SARS-CoV-2 infection for all vaccines were generally lower for Delta compared to the B.1.1.519 variant. Interestingly, effectiveness against COVID-19 related mortality was surprisingly lower for B.1.1.519, confirming previous data suggesting increased severity for cases infected with this variant, which comprised up to 89% of cases in Mexico by February 2021^39^.

We also report on a reduction in VE against SARS-CoV-2 infection and COVID-19 related hospitalization and death for most evaluated vaccines in older adults and persons living with diabetes. Previous data had shown that waning of vaccine-induced immunity was associated with increased age and comorbidity^42^. Our observations of reduced vaccines effectiveness in people living with diabetes also supports previous evidence which reported reduced neutralizing antibody response and immunogenicity were observed in people with diabetes^43^. These results are particularly relevant in Mexico, where diabetes is a leading cause of morbidity, increasing risk of severe COVID-19 and COVID-19 mortality^1,33,44^. Reductions in VE against SARS-CoV-2 infection were most pronounced for the ChAdOx1, Gam-COVID-Vac, CoronaVac and BNT162b2 vaccines, with more stability for the Ad26.COV2.S, Ad5-nCoV and mRNA-1273 vaccines both for older adults and persons living with diabetes. These results should be considered when evaluating prioritization and eligibility for vaccine boosters, which should be allocated to higher risk individuals.

Our study had some strengths and limitations. Amongst the strengths we highlight the use of a nation-wide surveillance system to monitor SARS-CoV-2 infections which can capture dynamics of COVID-19 transmission at the municipal level, allowing for evaluation of vaccination status and clinical follow-up of all suspected, confirmed, and negative COVID-19 cases in Mexico within the SISVER registry. This allowed us to evaluate three relevant outcomes for VE, including SARS-CoV-2 infection, and COVID-19 related hospitalization and death at a national level^27^. Furthermore, by evaluating a national vaccination campaign which incorporated multiple vaccines we were able to assess the feasibility of incorporating multiple vaccines simultaneously to maximize vaccination coverage, which will also be helpful when evaluating policies to implement heterologous schemes for vaccine boosters in the future. Finally, by incorporating a time-varying component to the vaccination status, we were able to capture shifts in exposure risks through the follow-up time to allow for more precise estimates of vaccine effectiveness for all evaluated outcomes. We would also like to acknowledge some limitations which should be considered to adequately interpret our results. First, the SISVER dataset is a sentinel surveillance system, which is designed to evaluate high-risk symptomatic cases which are assessed and tested for COVID-19^31^; because of its design, SISVER primarily identifies cases with moderate to severe COVID-19, which may skew our observations towards more severe cases and underrepresent mild and asymptomatic infections, which represent most SARS-CoV-2 cases; therefore, we are unable to distinguish VE in symptomatic versus asymptomatic SARS-CoV-2 infections^45^. This may also lead to a healthcare seeking bias, whereby identification of cases who did not seek testing or in-hospital care may be underrepresented in out estimation^27^. Second, information regarding vaccination status and date was collected during epidemiological characterization of SARS-CoV-2 cases and relies on accurate reporting by cases, which precludes precise evaluation of vaccine effectiveness waning through time, which should be evaluated in further studies. Third, estimates of hospitalization should be taken with caution, particularly considering the effect of hospital saturation in admittance of COVID-19 cases in Mexico during the second and third waves of the COVID-19 pandemic, which are included in the study^3^. Fourth, because vaccination roll-out in Mexico has been sequential and based on age categories, representation of vaccinated cases under 29 years has been low in comparison to the incidence of COVID-19 in this age group, which is heavily represented in the unvaccinated group. Even though all our models adjusted for age, the possibility of residual confounding in our estimations cannot be formally ruled out. Finally, because of the period we evaluated in our study, we were unable to estimate effectiveness during periods of increased circulation of the Omicron SARS-CoV-2 variant in Mexico, which had reported increased immune evasion and reduced VE for infection in different settings^46,47^. Further evaluation is required to assess the impact of the Omicron variant in VE in Mexico and the potential role of vaccine boosters in increasing protection against infection and severe forms of COVID-19 associated with this variant, as well as the impact of previous infection in modifying VE^48^.

In conclusion, all nationally available vaccines in Mexico were protective against SARS-CoV-2 infection and COVID-19 related hospitalization and death. This is the first report to simultaneously evaluate the effectiveness of seven different vaccines and amongst the first population-based reports for the effectiveness of the Gam-COVID-Vac and Ad5-nCoV vaccines. VE against SARS-CoV-2 infection was lower in periods of predominant circulation of the Delta variant compared to periods of B.1.1.519 predominance; however, most vaccines were more protective against COVID-19 related death during periods of high Delta circulation compared to B.1.1.519. Similarly, VE against SARS-CoV-2 infection and COVID-19 related death were lower for adults ≥60 years and people living with diabetes for most evaluated vaccine products, which should be taken into consideration when evaluating eligibility for further vaccine rollout and booster allocation.

## Data Availability

Code and materials are available for reproducibility of results at https://github.com/oyaxbell/covid_vaccines/. Data is available upon request to the General Directorate of Epidemiology of the Mexican Ministry of Health.

## ACKNOWLEDGMENTS

This project was registered and approved by the Research Committee at Instituto Nacional de Geriatría, project number DI-PI-006/2020. NEAV and CAFM are enrolled at the PECEM Program of the Faculty of Medicine at UNAM. NEAV is supported by CONACyT.

## AUTHOR CONTRIBUTIONS

Research idea and study design: OYBC, NEAV, SIVF, CAFM, LFC, DRG LMGR; data acquisition: JAMG,SECL, SECL, CAZJ, GCS, GGR, RCA, HLG, LMGR; analysis/interpretation: OYBC, NEAV, SIVF, CAFM, LFC, DRG; statistical analysis: OYBC, NEAV; manuscript drafting: OYBC, NEAV, CAFM, LFC, DRG, SIVF, LMGR, JMG, AKG, JAMG,SECL, HLG, LMGR, JAAF, CHZG, GRT; supervision or mentorship: OYBC, LMGR, SECL, CAZJ. Each author contributed important intellectual content during manuscript drafting or revision and accepts accountability for the overall work by ensuring that questions pertaining to the accuracy or integrity of any portion of the work are appropriately investigated and resolved.

## DISCLOSURES

SECL, GCS, CAZJ, GGR, RCA and HLG work at the Mexican Ministry of Health and oversee monitoring SARS-Cov-2 epidemiological surveillance and designing COVID-19 vaccination policies in Mexico. Data analysis and interpretation was primarily conducted by members of the team independent of these duties.

## FUNDING

This research was supported by Instituto Nacional de Geriatría.

## DECLARATIONS

All proceedings and evaluations for the use of anonymized data were approved by the Research and Ethics committee at Instituto Nacional de Geriatría, with approval number DI-PI-006/2020

## Notes

### Funding Statement

This research was supported by Instituto Nacional de Geriatria.

### Author Declarations

All proceedings and evaluations for the use of restricted anonymized data were approved by the Research and Ethics committee at Instituto Nacional de Geriatria, with approval number DI-PI-006/2020

### Summary of Updates

This version is updated after comments from reviewers.

