## Supplementary Material for "Effectiveness of a nation-wide COVID-19 vaccination program in Mexico"

**SUPPLEMENTARY MATERIAL – Effectiveness of a nation-wide COVID-19 vaccination program in Mexico**

**SUPPLEMENTARY TABLES**

**Supplementary Table 1.** Subjects evaluated for suspected COVID-19 in the nation-wide SISVER registry in Mexico rom December 24^th^, 2020 until September 27^th^, 2021 comparing by vaccination status and vaccine product

| Variable | Unvaccinated  (n=4792338) | ChAdOx1  (n=292800) | BNT162b2  (n=250806) | Ad5-nCoV  (n=87585) | Gam-COVID-Vac  (n=79551) | CoronaVac  (n=62474) | Ad26.COV2.S  (n=14870) | mRNA-12732  (n=5401) |
| --- | --- | --- | --- | --- | --- | --- | --- | --- |
| Follow-up (person-days) | 549,685,635 | 67,843,668 | 54,546,760 | 19,993,723 | 18,019,736 | 14,234,108 | 3,432,393 | 1,183,357 |
| Age (years, Median, IQR) | 37 (27-50) | 43 (35-49) | 46 (33-57) | 41 (33-50) | 52 (34-59) | 47 (32-64) | 30 (24-38) | 44 (24-57) |
| Males (%) | 2487088 (51.9) | 163210 (55.7) | 152921 (61.0) | 55663 (63.7) | 45819 (57.6) | 34568 (55.3) | 8222 (55.3) | 2881 (55.3) |
| 18-29 years (%) | 1515115 (31.6) | 30499 (10.4) | 44405 (17.7) | 14236 (16.3) | 10618 (13.3) | 11935 (19.1) | 6868 (46.2) | 1374 (25.4) |
| 30-59 years (%) | 2722383 (56.8) | 225942 (77.2) | 158707 (63.2) | 64874 (74.1) | 49124 (61.75) | 28765 (46.0) | 7664 (51.5) | 2872 (53.2) |
| >60 years (%) | 554840 (11.6) | 36359 (12.4) | 47694 (19.0) | 8475 (9.7) | 19809 (61.7) | 21774 (34.9) | 338 (2.3) | 1155 (21.4) |
| Fully vaccinated (%) | - | 60250 (20.6) | 197078 (78.8) | 87585 (100.0) | 39952 (50.2) | 34469 (55.2) | 14870 (100.0) | 3764 (69.7) |
| COVID-19 incidence (per 1,000) | 2.672 | 1.440 | 1.038 | 1.577 | 0.868 | 1.803 | 0.978 | 0.489 |
| Hospitalization (per 1,000) | 0.383 | 0.111 | 0.090 | 0.164 | 0.048 | 0.260 | 0.054 | 0.048 |
| Death (per 1,000) | 0.191 | 0.039 | 0.031 | 0.059 | 0.019 | 0.110 | 0.011 | 0.009 |
| Diabetes (%) | 363864 (7.6) | 28609 (9.8) | 30160 (12.0) | 8371 (9.6) | 9395 (11.8) | 9446 (15.1) | 626 (4.2) | 453 (8.4) |
| Obesity (%) | 423535 (8.8) | 30964 (10.5) | 28515 (11.4) | 9712 (11.2) | 5638 (8.22) | 7363 (11.8) | 1717 (11.5) | 385 (7.1) |
| CKD (%) | 44378 (0.9) | 2344 (0.8) | 2460 (1.0) | 670 (0.8) | 555 (0.7) | 814 (1.3) | 31 (0.2) | 19 (0.4) |
| COPD (%) | 29114 (0.6) | 1847 (0.6) | 2256 (0.9) | 623 (0.7) | 608 (0.7) | 874 (7.6) | 37 (0.2) | 27 (0.5) |

**Abbreviations:** CKD, Chronic Kidney Disease; COPD, Chronic Obstructive Pulmonary Disease

**Supplementary Table 2.** Description of all nationally available COVID-19 vaccine products authorized for emergency use in Mexico.

| **Vaccine product** | **Mechanism of action** | **Doses for fully vaccinated** | **Regulatory status in Mexico** | **Authorization date** |
| --- | --- | --- | --- | --- |
| BNT162b2  (Pfizer, Inc,/BioNTech) | mRNA vaccine | 2 | Authorized for emergency use in adults | 11/December/2020 |
| ChAdOx1  (AstraZeneca) | Non-repplicant adenovirus vaccine | 2 | Authorized for emergency use in adults | 04/01/2021 |
| Gam-COVID-Vac  (Sputnik-V) | Non-replicant adenovirus vaccine | 2 | Authorized for emergency use in adults | 02/02/2021 |
| CoronaVac (Sinovac) | Inactivated virus vaccine | 2 | Authorized for emergency use in adults | 09/02/2021 |
| Ad5-nCoV (CanSino Biologics) | Non-replicant adenovirus vaccine | 1 | Authorized for emergency use in adults | 09/02/2021 |
| Ad26.COV2-S  (Johnson&Jonson) | Non-replicant adenovirus vaccine | 1 | Authorized for emergency use in adults | 27/05/2021 |
| mRNA-12732 (Moderna) | mRNA vaccine | 2 | Authorized for emergency use in adults | 17/08/2021 |

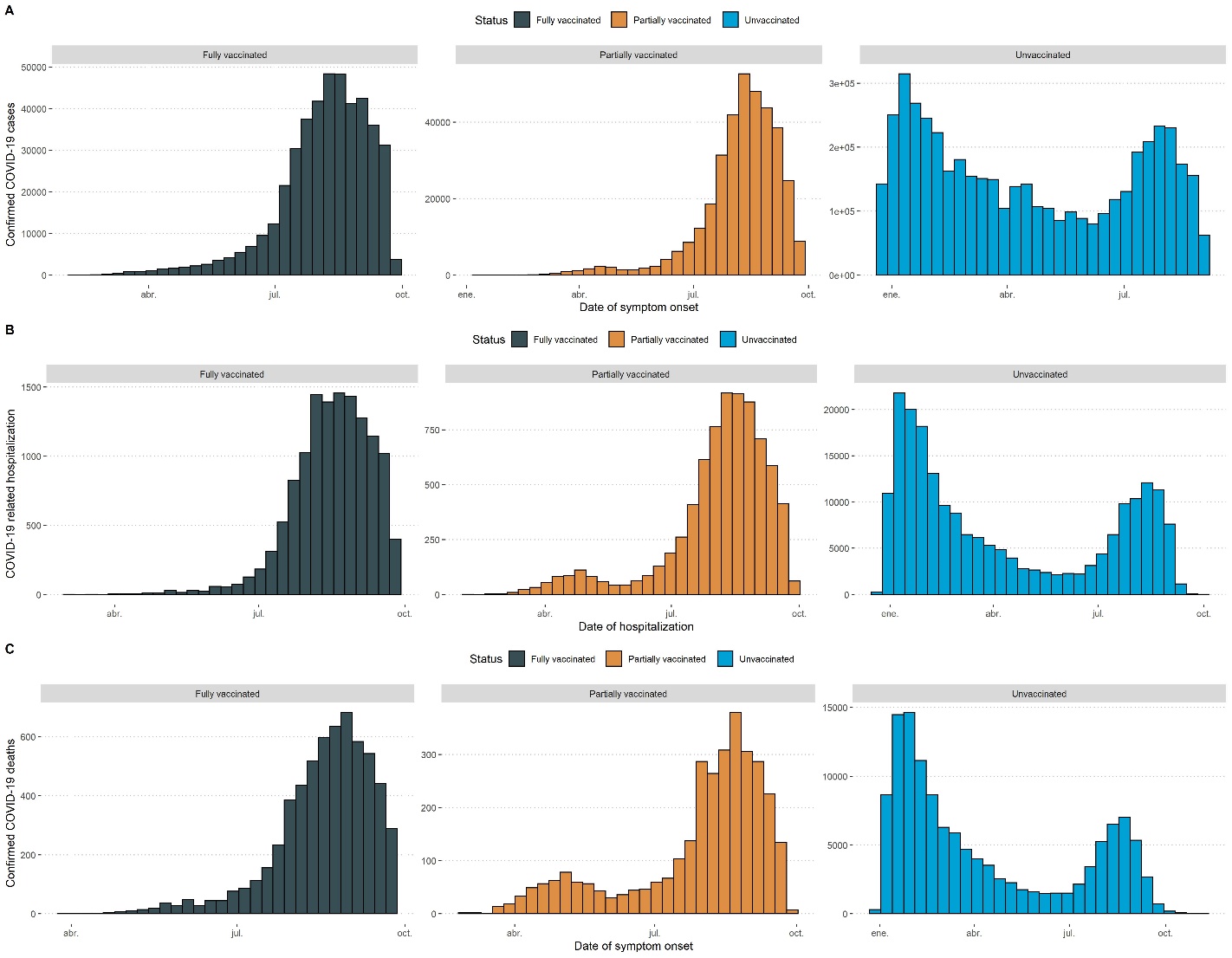

**Supplementary Figure 1.** Incidence of symptomatic, laboratory confirmed COVID-19 in Mexico stratified by vaccination status (A), as well as incidence of COVID-19 related hospitalization (B) and deaths (C).

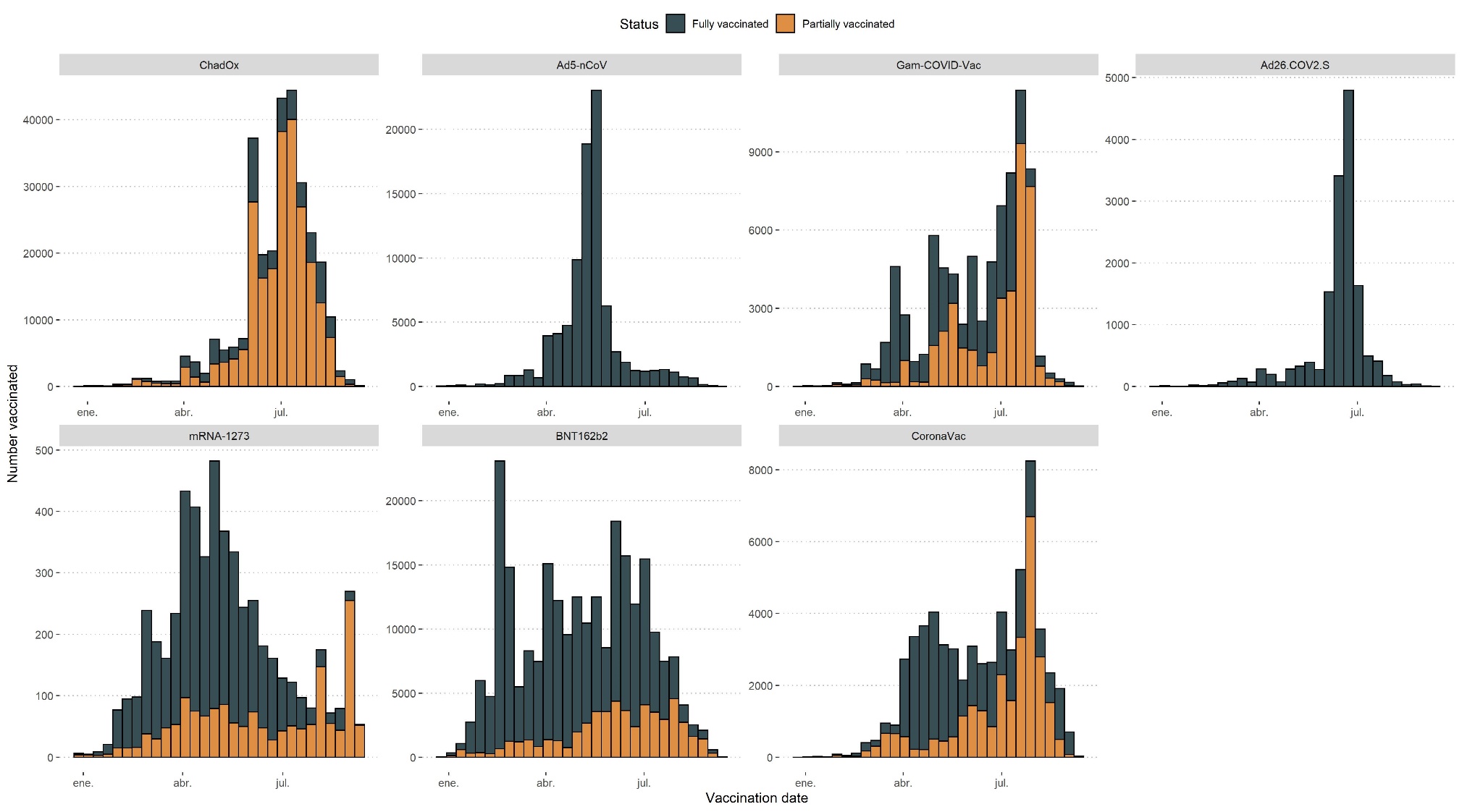

**Supplementary Figure 2.** Vaccination date of cases evaluated in the SISVER platform stratified by individual vaccine product implemented within the Mexican National vaccination program.

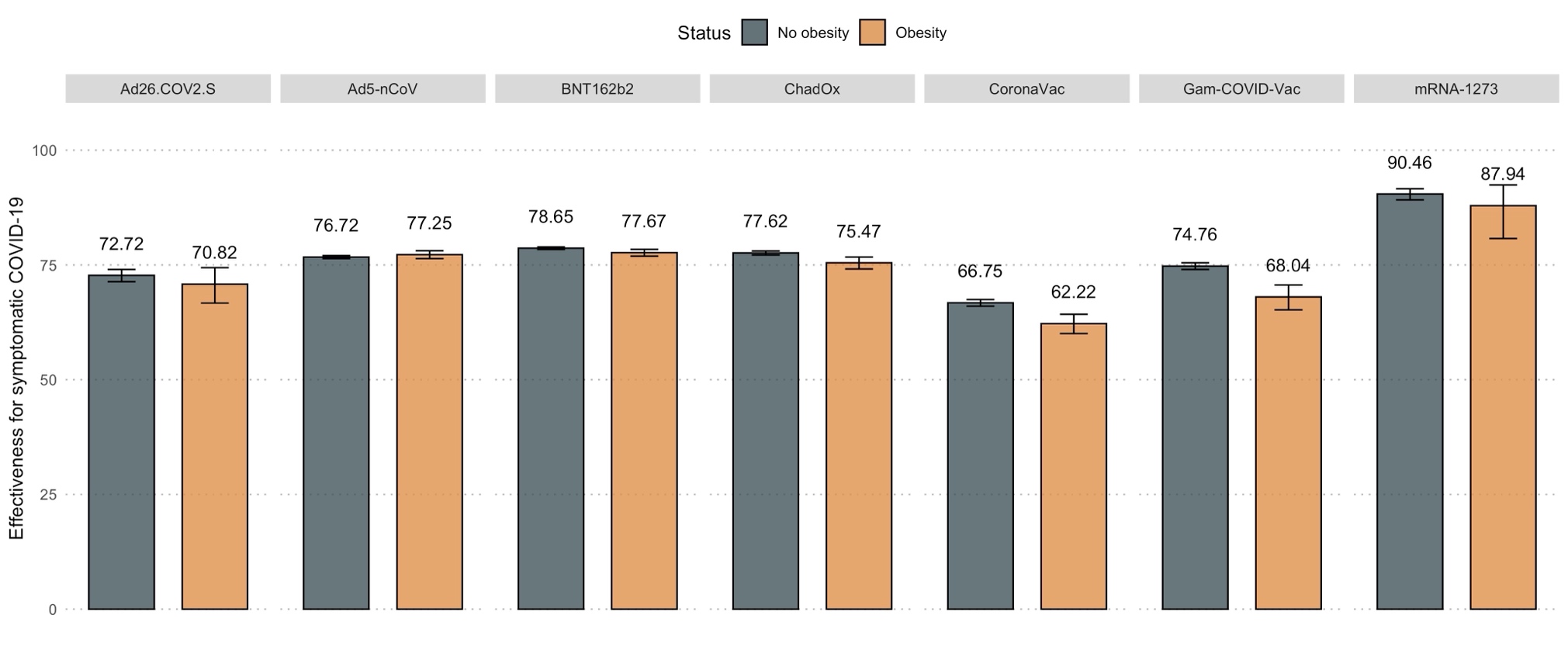
 **Supplementary Figure 3.** Vaccine effectiveness against symptomatic laboratory confirmed SARS-CoV-2 infection for all nationally available COVID-19 vaccine products in Mexico, comparing individuals with and without obesity.

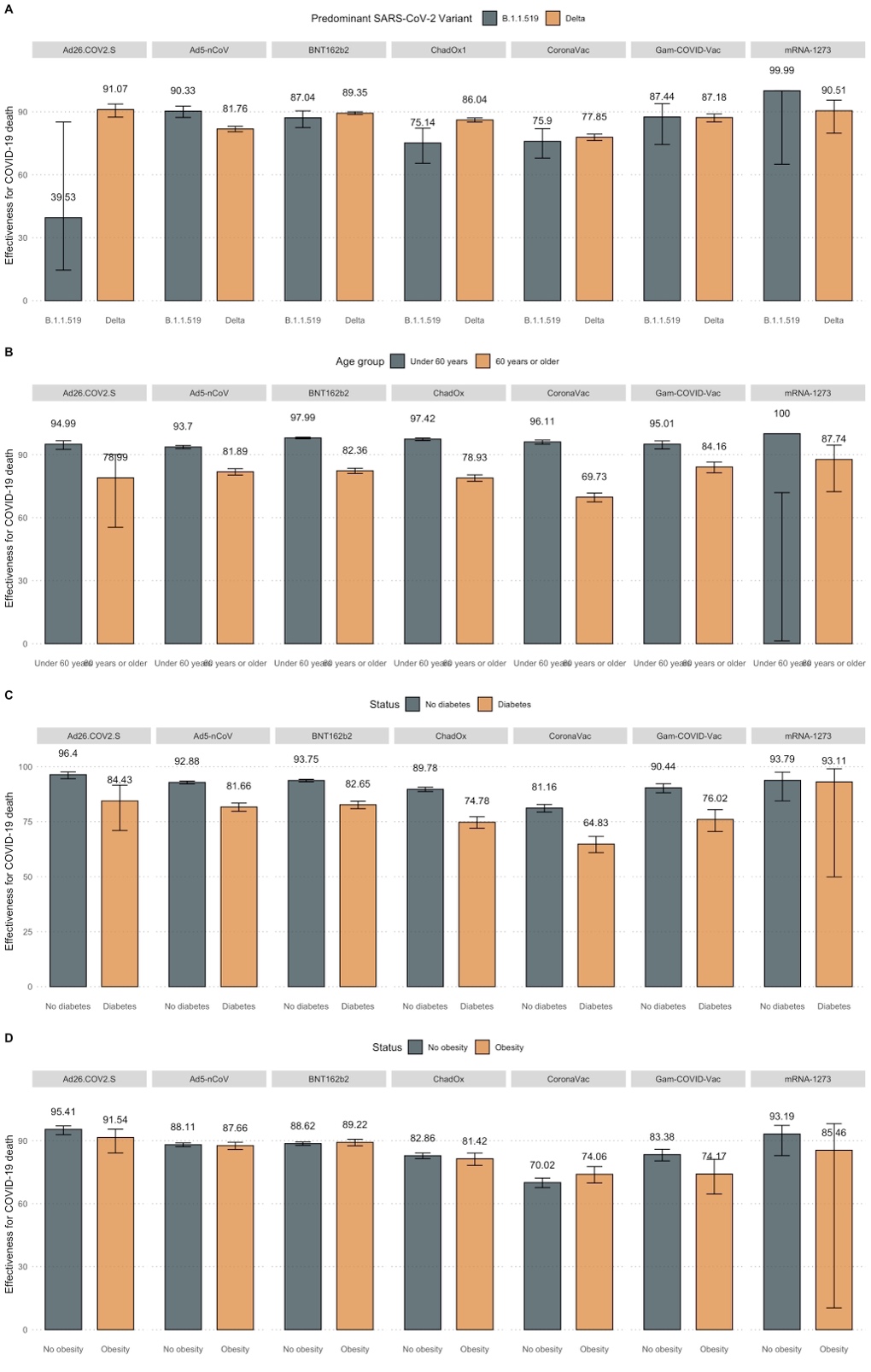

**Supplementary Figure 4.** Estimated vaccine effectiveness against COVID-19 related death for nationally available COVID-19 vaccines stratified by predominant circulating SARS-CoV-2 variant (B.1.1.519 vs. B.1.617.2, A), age group (over vs. under 60 years, B) and diabetes (C) or obesity status (D).
